# Distributing Safer Smoking Pipes Increases Engagement with Harm Reduction Services in the United States: Findings from the National Survey of Syringe Services Programs

**DOI:** 10.1101/2024.06.28.24309683

**Authors:** Esther O. Chung, Sheila V. Patel, Lynn D. Wenger, Jamie L. Humphrey, Ricky N. Bluthenthal, Hansel E. Tookes, Don C. Des Jarlais, Sara N. Glick, Paul A. LaKosky, Stephanie Prohaska, Laura Guzman, Alex H. Kral, Barrot H. Lambdin, National Survey of Syringe Services Programs Group

## Abstract

**Background:** In response to the recent and growing shift from injecting opioids to smoking fentanyl, an increasing number of syringe services programs (SSPs) in the USA are distributing safer smoking supplies. A recent federal ban prevents SSPs from using federal funding to procure safer smoking supplies. There is a lack of research on safer smoking supply distribution and harm reduction outcomes. Therefore, we assessed the relationship between distribution of safer smoking supplies by SSPs and levels of participant engagement and naloxone distribution.

**Methods:** We used data from the 2023 National Survey of Syringe Services Programs (NSSSP) (N=429), which measured services delivered in 2022. SSPs reported whether they distributed safer smoking pipes (yes/no). We examined the relationship between safer smoking pipe distribution and two outcomes: the number of participant encounters and naloxone doses distributed.

**Results:** There were 187 SSPs (43.6%) that distributed pipes for smoking to participants. Compared to SSPs that did not distribute pipes, those that distributed pipes reported more participant encounters (aRR=1.49, 95% CI: 1.09-2.02) and naloxone dose distribution (aRR=1.21, 95% CI: 0.89-1.65), though this latter finding was not statistically significant.

**Conclusions:** Findings showed SSPs distributing safer smoking pipes had more participant engagement and naloxone distribution. To maximize their full individual and population-level health benefits, SSPs should be supported technically, legally, and financially to implement safer smoking supply distribution for their participants.

**Highlights:**

- In 2022, 44% of syringe services programs (SSPs) distributed safer smoking pipes.
- More community-based organizations distributed pipes than public health or health care programs.
- Distribution of safer smoking pipes was associated with more participant encounters.

## 1. Introduction

For the past two decades, the overdose mortality rate in the United States (U.S.) has steadily increased (Ahmad et al., 2023). Since 1999, over one million people have died of a drug overdose (Centers for Disease Control and Prevention, 2023). In 2022, there were over 100,000 drug overdose fatalities and opioids were involved in over 75% of overdose deaths (Ahmad et al., 2024). Since 2015, synthetic opioids, including fentanyl and fentanyl analogs, have comprised the majority of opioid overdose deaths, while deaths due to heroin have decreased (Spencer et al., 2023). Furthermore, in 2022, roughly 1 in 14 HIV diagnoses in the U.S. were attributable to injection drug use (Centers for Disease Control and Prevention, 2024b), and the estimated rate of new hepatitis C virus (HCV) cases among people who inject drugs was 2.2 per 100,000 population (Centers for Disease Control and Prevention, 2024a).

Syringe services programs (SSP) are proven, effective prevention programs that distribute safe drug use supplies, such as new or sterile needles, smoking supplies, and other equipment, with the goal of reducing the health related harms associated with drug use (Paone et al., 1995; Bluthenthal et al., 2000; Aspinall et al., 2014; Jakubowski et al., 2023). In addition to providing safe drug use supplies, SSPs often provide other evidence-based interventions, such as overdose education and naloxone distribution (OEND), infectious disease testing and management, and linkage to other healthcare and social services. Naloxone is a critical, life-saving intervention that is highly effective at reversing opioid overdoses. Since 1996, SSPs have led efforts to provide evidence-based OEND programs (Wheeler et al., 2012, 2015; Bennett & Elliott, 2021) that train laypeople, including people who use drugs (PWUD), family members, and peers, to respond to overdose events.

Recent studies have documented changes from injecting to smoking drugs, particularly along the West Coast of North America, as illegally-manufactured fentanyl entered the unregulated drug market (Kral et al., 2021; Parent et al., 2021; Kingston et al., 2022; Megerian et al., 2024; Eger et al., 2024). PWUD report preferring smoking over injecting substances for a variety of reasons, including lower financial costs of fentanyl compared to heroin, reduced stigma, improved health benefits (e.g. fewer skin and soft tissue infections), and perceived lower risk of overdose compared to injecting (Kral et al., 2021; Ivsins et al., 2024). Secondary analyses of mortality records between 2020 and 2022 identified a marked increase in overdose deaths with evidence of smoking and a decrease in deaths with evidence of injection (Tanz et al., 2024).

However, as this study did not compare the risks of fatal overdose between people who inject and people who smoke, these findings likely reflect broader drug use trends. A recent study in California that compared risks of non-fatal overdose found a 40% increased prevalence among people who inject fentanyl compared to people who smoke fentanyl (Megerian et al., 2024). The recent transition to smoking is not limited to those using opioids. People who use stimulants, such as methamphetamine and cocaine, have also reported switching to smoking, primarily due to overdose fears when non-prescribed fentanyl entered the unregulated drug market (Ivsins et al., 2024). As the prevalence of polysubstance use involving fentanyl and methamphetamine rises (Friedman & Shover, 2023; Han et al., 2021), it is crucial that service organizations adapt to the growing needs of PWUD, which involves access to harm reduction and treatment.

Prior research has shown providing safer smoking pipes can lead to reductions in the frequency of injections and risky smoking practices (e.g. pipe sharing, using broken pipes), as well as improved health outcomes among those who smoke drugs, such as fewer burns and cuts (Leonard et al., 2008; Bridge, 2010; Stöver & Schäffer, 2014; Frankeberger et al., 2019; Dunleavy et al., 2021; Fitzpatrick et al., 2022; Tapper et al., 2023). Given SSPs are trusted by PWUD (MacNeil & Pauly, 2011), they are ideal programs for distributing safer smoking supplies. Engaging PWUD through the distribution of safer smoking supplies at SSPs may also increase access to other evidence-based interventions, such as naloxone. Combined HIV and HCV prevention interventions with syringe distribution were associated with reduced transmission, risky behaviors, and other adverse outcomes (Stopka et al., 2007; Des Jarlais et al., 2014; Uusküla et al., 2015; Platt et al., 2018). Extrapolating from this literature, it is plausible that combining safer smoking supplies with naloxone distribution may reduce overdose rates among SSP participants. However, SSPs that only provide safe injection supplies and not smoking supplies may not meet the needs of a key subpopulation of people who smoke drugs. While some SSPs have provided safer smoking supplies (e.g. new glass pipes, aluminum foil, pipe covers) for years, more SSPs started providing such supplies in response to the shift towards smoking drugs (Fitzpatrick et al., 2022; Singh et al., 2022; Office of AIDS, n.d.). Emerging research from Seattle, Washington show a high interest and need for safer smoking supplies among SSP participants (Reid et al., 2023; Kingston et al., 2024). However, there remains a dearth of research on safer smoking supply distribution in other parts of the U.S., especially within the context of people transitioning from injection to smoking.

To address these gaps in the literature, we aimed to assess the relationship of the distribution of safer smoking pipes by SSPs with participant engagement and naloxone distribution using the National Survey of Syringe Services Programs (NSSSP). We hypothesized pipe distribution would be associated with an increased number of participant contacts and naloxone doses distributed by SSPs.

## 2. Methods

### 2.1. Data source

Our primary data source was the 2023 NSSSP (Lambdin, Patel, et al., 2023), an annual cross-sectional survey fielded by RTI International to understand the health and social services delivered by SSPs. SSPs were defined as community-based programs that, at minimum, provide free drug use supplies (e.g., new or sterile syringes, smoking equipment, and other safe use supplies) to PWUD with the goal of reducing harms associated with substance use. SSPs often provide other health and social services, such as drug checking, naloxone distribution, access to medications for opioid use disorder, and linkage to other medical care and social support services. RTI International and the North American Syringe Exchange Network (NASEN) collaborated to develop the full sampling frame of U.S. SSPs (Lambdin, Patel, et al., 2023; Patel et al., n.d.). NASEN’s dataset included SSPs that agreed to be publicly listed in their directory; SSPs that did not wish to be publicly listed but agreed to be contacted for research purposes; SSPs that belonged to NASEN’s Buyer’s Club that were not part of the directory; and other known SSPs that were not in NASEN’s directory or did not participate in their Buyer’s Club. RTI’s dataset included SSPs that responded to prior NSSSPs and SSPs that were proactively identified through SSP networks and partner agencies, including the National Harm Reduction Coalition and state health departments. RTI and NASEN merged their datasets of known SSPs, removed duplicates, and identified non-operational SSPs.

For the present study, we used data from the 2023 NSSSP, which asked about services delivered in 2022. All SSPs known to be operating in 2022 were invited to participate in an online survey between March and August 2023 and received $125 as compensation. Respondents provided information about their SSP in 2022 on the following topics: organizational characteristics, community support and opposition, drug use supply distribution, overdose prevention, and other service provision among other topics. Out of the 626 SSPs known to be operating in 2022, 471 (75%) responded to the 2023 NSSSP survey.

### 2.2. Measures

Our exposure of interest was whether an SSP distributed pipes to participants for smoking in 2022. Specifically, respondents were asked “For each of the following drug use supplies (other than syringes), please indicate which ones were provided to participants in 2022. Please check all that apply.” The list of supplies included pipes for smoking. We examined two outcomes: number of participant encounters and number of naloxone doses provided. For participant contacts, respondents were asked “How many participant contacts occurred at your SSP in 2022? By participant contacts, we mean the number of encounters / participant visits made to your SSP during 2022.” For naloxone doses, respondents were asked “How many naloxone doses did your syringe services program distribute in 2022?”

Informed by findings from prior research (Lambdin et al., 2020; Facente et al., 2024; Ray et al., 2024), we selected the following covariates *a priori* to include in our models: annual budget of the SSP, organizational type, urbanicity, opioid overdose mortality rates, and U.S. regional Census division. Annual SSP program budget was classified into the following categories: <$50,000, $50,000 - $149,999, $150,000 - $499,999, $500,000 - $999,999, and over $1,000,000. Due to sample size restrictions, we further categorized SSP budget into < $150,000 vs. ≥ $150,000 to obtain relatively even groups. We classified SSP organizational type based on community-based organization (CBO) vs. another type of organization, such as departments of public health (DPH), health care organizations (HCO), or other. Using the National Center for Health Statistics (NCHS) Urban-Rural Classification Scheme and guidance from the Pew Research Center (Parker et al., 2018), we constructed a county-level measure of urbanicity with three categories: urban, suburban, and rural based on the SSP’s headquarters. We also used 2020 opioid overdose mortality rates per 100,000 from the National Vital Statistics System as a proxy measure for the level of need in the community (Rossen et al., 2022). We standardized mortality rates with a mean of zero and standard deviation of one. Finally, we classified each SSP into regional U.S. Census divisions (New England, Middle Atlantic, East North Central, West North Central, South Atlantic, East South Central, West South Central, Mountain, and Pacific). SSPs operating in Puerto Rico were grouped with South Atlantic.

### 2.3. Statistical analyses

We first summarized the data using descriptive statistics. Using negative binomial models, we then estimated adjusted rate ratios (aRR) and 95% confidence intervals (CI) of the total effect of pipe distribution on number of participant encounters and naloxone doses distributed. We adjusted for annual budget, organization type, urbanicity, and opioid overdose mortality rates. We included fixed effects for regional census divisions to account for clustering.

We imputed missing data using multiple imputation by chained equations (MICE) (Van Buuren & Groothuis-Oudshoorn, 2011; White et al., 2011; Graham, 2009). We generated 50 datasets, including all analytic variables and auxiliary variables (see Appendix Methods for further details). Based on prior recommendations to improve model precision (Von Hippel, 2007; White et al., 2011), we restricted the analytic sample to SSPs with complete information with regards to our study outcomes (n=429). Then, we ran multivariable models within each imputed dataset, and pooled results across all datasets. Statistical analyses were conducted using Stata 18 (College Station, TX) and R, version 4.3.2 (R Core Team, 2023).

## 3. Results

### 3.1. Sample characteristics

Across our analytic sample (Table 1, n=429), the median annual number of participant contacts was 1,400 (Interquartile range (IQR): 260 - 4,100) and the median annual number of distributed naloxone doses was 2,120 (IQR: 507 - 6,000). The majority of SSPs were classified as CBOs (57.6%). Nearly half of SSPs operated on an annual budget less than $150,000. The majority of SSPs were headquartered in suburban areas (47.2%). Our sample included SSPs in each of the nine U.S. Census divisions, with the greatest number concentrated in the Mountain (20.5%), Pacific (18.6%), and South Atlantic (17.5%) regions.

**Table 1.** Sample characteristics, National Survey of Syringe Services Programs, 2023^a^

|  | Overall<br>(N=429) | Pipe distribution<sup>b</sup> |  |
| --- | --- | --- | --- |
|  |  | Distributed<br>pipes<br>(N=187) | Did not distribute<br>pipes<br>(N=240) |
| **Participant contacts** |  |  |  |
| Median [Q1 - Q3] | 1,400<br>[260 - 4,100] | 2,490<br>[797 - 7,080] | 867<br>[134 - 2,370] |
| **Naloxone doses distributed** |  |  |  |
| Median [Q1 - Q3] | 2,120<br>[507 - 6,000] | 4,110<br>[1,750 - 9,960] | 1,000<br>[185 - 4,000] |
| **SSP type** |  |  |  |
| CBO | 247 (57.6%) | 149 (79.7%) | 97 (40.4%) |
| DPH | 134 (31.2%) | 22 (11.8%) | 112 (46.7%) |
| HCO/Other | 44 (10.3%) | 16 (8.6%) | 27 (11.3%) |
| Missing | 4 (0.9%) | 0 (0%) | 4 (1.7%) |
| **Total annual budget** |  |  |  |
| < \$50,000 | 84 (19.6%) | 38 (20.3%) | 45 (18.8%) |
| \$50,000 - \$149,999 | 69 (16.1%) | 28 (15.0%) | 40 (16.7%) |
| \$150k - \$499,999 | 89 (20.7%) | 48 (25.7%) | 41 (17.1%) |
| \$500k - \$999,999 | 32 (7.5%) | 21 (11.2%) | 11 (4.6%) |
| > \$1 million | 26 (6.1%) | 18 (9.6%) | 8 (3.3%) |
| Missing | 129 (30.1%) | 34 (18.2%) | 95 (39.6%) |
| **Urbanicity** |  |  |  |
| Suburban | 203 (47.3%) | 85 (45.5%) | 118 (49.2%) |
| Rural | 104 (24.2%) | 27 (14.4%) | 77 (32.1%) |
| Urban | 111 (25.9%) | 72 (38.5%) | 39 (16.3%) |
| Missing | 11 (2.6%) | 3 (1.6%) | 6 (2.5%) |
| **Opioid overdose death rate per 100,000 (2020)** |  |  |  |
| Median [Q1-Q3] | 30.8 [22.8 - 41.1] | 29.0 [21.6 - 38.4] | 31.7 [23.5 - 45.0] |
| Missing | 12 (2.8%) | 3 (1.6%) | 7 (2.9%) |
| **Census divisions** |  |  |  |
| Pacific | 80 (18.6%) | 50 (26.7%) | 29 (12.1%) |
| New England | 29 (6.8%) | 22 (11.8%) | 7 (2.9%) |
| Middle Atlantic | 26 (6.1%) | 17 (9.1%) | 9 (3.8%) |
| East North Central | 67 (15.6%) | 31 (16.6%) | 36 (15.0%) |
| West North Central | 21 (4.9%) | 13 (7.0%) | 8 (3.3%) |
| South Atlantic | 75 (17.5%) | 28 (15.0%) | 46 (19.2%) |
| East South Central | 29 (6.8%) | 3 (1.6%) | 26 (10.8%) |
| West South Central | 14 (3.3%) | 8 (4.3%) | 6 (2.5%) |
| Mountain | 88 (20.5%) | 15 (8.0%) | 73 (30.4%) |

<sup>a</sup> We present summary statistics that were observed prior to imputation. For multivariable analyses, we imputed values for missing variables using multiple imputation. The overall sample was restricted to SSPs that were not missing information on outcomes.
<sup>b</sup> Two SSPs were missing information on pipe distribution and were not included in the stratification by pipe distribution.
Abbreviations: Quartile 1 (Q1); Quartile 3 (Q3); Syringe services program (SSP); Community-based organization (CBO); Department of Public Health (DPH); Health care organization (HCO)

A total of 187 SSPs (43.6%) reported distributing pipes to participants. A greater number of CBO SSPs distributed pipes compared to DPH or HCO/Other (79.7% vs. 11.8% and 8.6%, respectively). Among the nine Census divisions, SSPs in the Pacific region had the greatest proportion that distributed pipes (26.7%).

### 3.2. Associations between pipe distribution and SSP services

SSPs that distributed pipes had a higher median number of participant encounters and naloxone doses distributed than those that did not distribute pipes (Table 1). SSPs that distributed pipes reported more participant encounters, compared to SSPs that did not distribute pipes (Figure 1, aRR=1.49, 95% CI: 1.09-2.02), after adjusting for SSP organizational type, budget, urbanicity, overdose mortality rates in 2020, and regional Census divisions. Furthermore, we found SSPs that distributed pipes reported more naloxone dose distribution than those that did not distribute pipes (Figure 1, aRR=1.21, 95% CI: 0.89-1.65), though this finding was not statistically significant.

**Figure 1.**
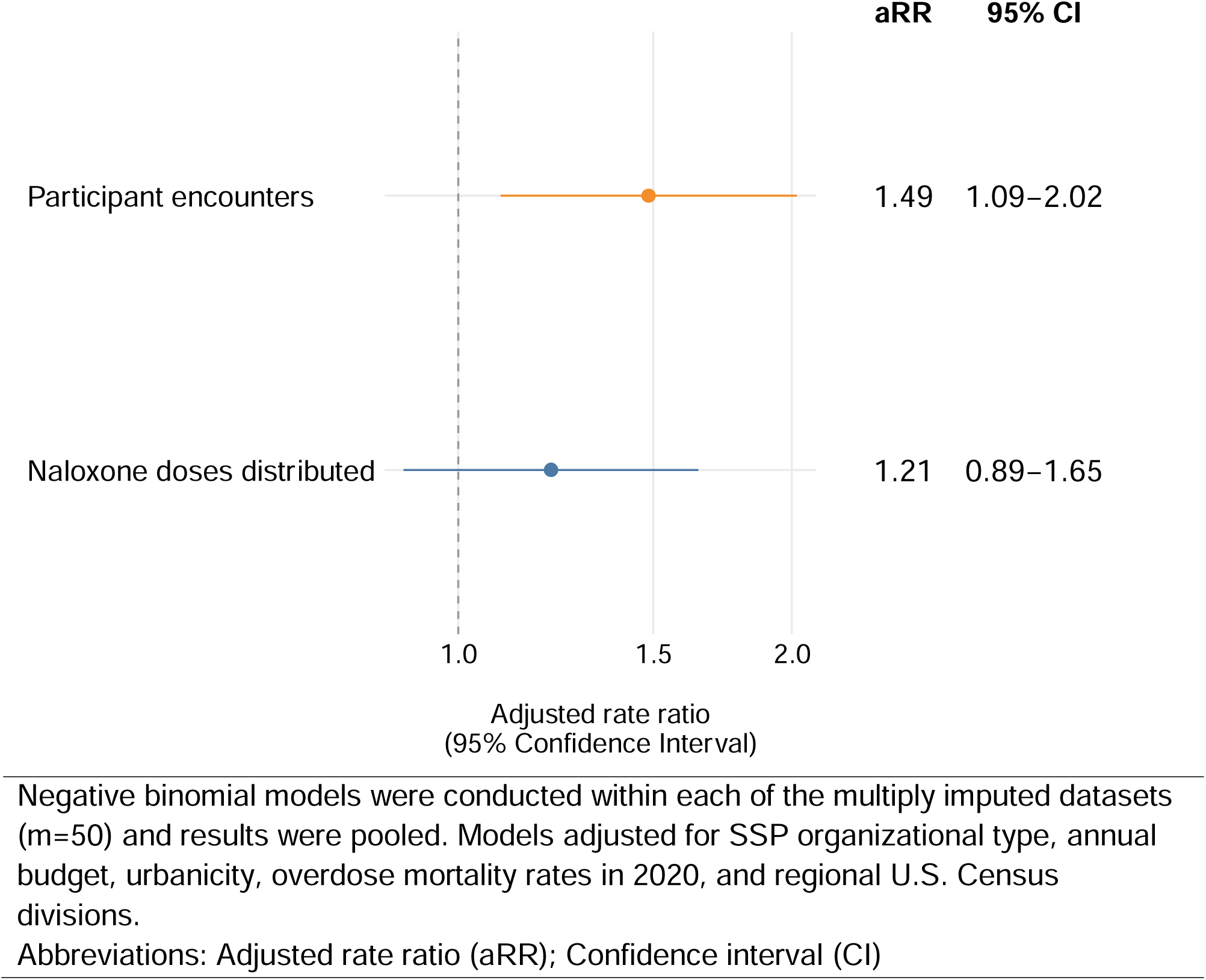
Associations between pipe distribution and participant encounters and naloxone distribution, NSSSP 2023 survey

We also ran two sensitivity analyses: (1) limiting to only complete cases (n=291) and (2) using the fully imputed dataset (n=471). Results from sensitivity analyses did not qualitatively change our findings (Appendix).

## 4. Discussion

Using data from the 2023 NSSSP, the largest sample of U.S. SSPs collected to date, we found 44% of SSPs reported distributing pipes for smoking. SSPs that offered pipes to their participants had a 49% higher rate of participant encounters compared to SSPs that did not offer pipes to their participants. Given SSPs can reduce transmission of HIV and hepatitis C virus (Fernandes et al., 2017; Platt et al., 2017), reduce overdose fatalities (Lambdin et al., 2020), and improve entry into substance use disorder treatment or clinical care (Jakubowski et al., 2023), implementation of pipe distribution by SSPs represents a critical approach that could increase participant engagement and reach a broader population of PWUD at risk for infectious disease or overdose. Since SSPs are trusted by PWUD (MacNeil & Pauly, 2011), it follows that bringing more people to SSPs more often has the potential to improve population-level health benefits of this critical public health approach. Further, this approach may be especially important in the era of fentanyl, as recent research suggests people who smoke fentanyl experience non-fatal overdoses, infectious diseases, and hospitalization less often than people who inject fentanyl (Megerian et al., 2024). SSPs offering pipescould help people adopt safer drug use practices by facilitating a transition from injection to smoking. Future studies should explore the implementation adjustments when SSPs offer smoking supplies, such as how they prepare for a higher volume of participant encounters through increased staffing and the consideration of neighborhood and community dynamics.

Research has shown a shifting trend in drug use patterns from injection toward smoking since 2020 (Kral et al., 2021; Eger et al., 2024). Some SSPs were providing smoking supplies prior to this shift, while others have more recently implemented safer smoking supply distribution to meet emerging needs and maintain engagement with their participants. However, access to and distribution of safer smoking supplies remain controversial, especially the distribution of smoking pipes. U.S. federal law prohibits the use of federal funds to purchase safer smoking supplies (Centers for Disease Control and Prevention, 2024c; US Department of Health and Human Services, 2016). The introduction of safer smoking supplies by SSPs was met with considerable public opposition, including the Cutting Rampant Access to Crack Kits (CRACK) Act by Senator Marco Rubio (R-FL) and other Republicans and the Preventing Illicit Paraphernalia for Exchange Systems (PIPES) Act by Senators Rubio and Manchin (D-WV) in February 2022 (Rubio, 2022a, 2022b). This public opposition culminated in the Biden administration revising its policies and disallowing SSPs from using federal funding for harm reduction to procure smoking supplies (Becerra & Gupta, 2022). In contrast to the public response to prescription opioid use primarily among White people (Netherland & Hansen, 2017), the political opposition against the distribution of smoking pipes appears to be a negative externality of the racially motivated War on Drugs, which has primarily targeted Black and Brown communities in the U.S. for the past 50 years (Cohen et al., 2022; Hagle et al., 2021). The federal ban and political opposition have forced SSPs that are distributing safer smoking supplies to patch together different funding sources—individual donations, private funding, state grants, etc.—to support implementation. SSP funding levels already do not meet minimum benchmarks, and insufficient funding threatens the implementation of evidence-based interventions (Akiba et al., 2024; Facente et al., 2024; Ray et al., 2024). Our findings highlight how critical it is to financially support SSPs to adapt to the evolving context and implement safer drug use supplies.

In our sample, SSPs classified as CBOs distributed pipes for smoking more often than SSPs classified as DPHs or HCOs. This finding aligns with recent research which found CBO SSPs provided a greater number of syringes, naloxone doses, fentanyl test strips, and buprenorphine compared to DPH SSPs (Ray et al., 2024). Given the federal ban on purchasing pipes, DPH SSPs may experience greater legal and financial obstacles for pipe distribution compared to CBO SSPs. CBO SSPs tend to adopt innovative approaches more quickly than DPHs or HCOs (Wenger et al., 2021).

Examples include expanding mail-based harm reduction supplies, low-barrier buprenorphine, and vaccinations during the COVID-19 pandemic, and delivering HIV pre-exposure prophylaxis (PrEP) and buprenorphine (Wenger et al., 2021; Aronowitz et al., 2021; Behrends et al., 2022; Hood et al., 2020; Heidari et al., 2024; Bartholomew et al., 2022). We also found SSPs in the Pacific Census division had the highest proportion of pipe distribution. This finding likely reflects how SSPs responded to the shift to smoking fentanyl along the West coast as well as more political and legal support for harm reduction services compared to other parts of the U.S. For example, the distribution of safer smoking supplies by SSPs is legal in California and Washington (Office of AIDS, n.d.; Wash. Rev. Code § 69.50.4121., 2023).

While not statistically significant, we found SSPs that distributed pipes had a 21% higher rate of naloxone dose distribution than SSPs that did not distribute pipes. Prior studies have shown most overdoses are reversed with naloxone by laypeople, primarily PWUD (Kingston, 2021; Lambdin, Wenger, et al., 2023). Previous research in the U.S. and Scotland have demonstrated increases in naloxone distribution in communities with increased risk for overdose fatalities were associated with reductions in opioid overdose mortality at the community-level (Bird et al., 2015; Walley et al., 2013; Bird & McAuley, 2019; Naumann et al., 2019). Despite large expansions in OEND (Des Jarlais et al., 2015; Lambdin et al., 2020), there remain important implementation gaps in naloxone saturation at the community level (Lambdin et al., 2018, 2020). Previous research has shown that implementation strategies to improve naloxone distribution by SSPs can improve access among SSP participants (Patel et al., 2023). Distribution of safer smoking supplies could be another programmatic opportunity for SSPs to not only increase engagement with participants, but also to increase naloxone distribution to communities most likely to prevent an opioid overdose from becoming fatal.

Our findings have important policy implications. State paraphernalia laws, which criminalize possession and sale of items used to consume non-prescribed drugs, prevent many SSPs across the country from legally distributing safer drug use supplies, including syringes and smoking equipment, as well as drug checking kits including items such as fentanyl test strips. Advocates, supported by years of research, have called for the repeal of these paraphernalia laws in order to allow PWUD to access more comprehensive and culturally appropriate health and social services at SSPs (Davis et al., 2019; Singer & Heimowitz, 2022). SSPs have also experienced a recent surge in community and law enforcement harassment related to the distribution of smoking supplies (Ovalle, 2024). Policymakers, public health officials, and law enforcement agencies should be informed about the emerging evidence showing that transitioning people from injection to smoking could have positive health benefits (Megerian et al., 2024). Our study adds to this body of research demonstrating that providing pipes for smoking increases participant engagement with evidence-based SSPs and naloxone distribution from SSPs. Based on our findings, policymakers should create supportive legal and funding environments for implementing safer drug use supplies so that community opposition and law enforcement do not compromise the full population-level benefits of SSP services.

Our findings should be interpreted in light of potential methodological limitations. First, this was a cross-sectional survey, representing a snapshot in time, and our exposure was not randomized between our two comparison groups. While we attempted to adjust for possible confounders, the potential for residual confounding from unmeasured or mismeasured variables remain. Future longitudinal studies would be valuable to understand changes over time before and after implementation of safer smoking supply distribution. Second, it is possible SSPs that did not respond to the survey could be less resourced and in unsanctioned environments, meaning experiences from these types of SSPs could be underrepresented. However, our overall response rate of 75% is very robust, and responding SSPs were diverse in terms of geographical region, budget, organizational type, and areas served. A third potential limitation is that some data were missing, which can lead to selection bias. We used multiple imputation to impute missing values and minimize the potential for such bias. Fourth, NSSSP data are self-reported by SSP staff, often with the assistance of programmatic records; however, responses are still subject to recall bias. Lastly, the 2023 NSSSP only asked whether SSPs distributed pipes to participants and did not ask about the types of pipes or other smoking supplies, such as foil, or the amount of smoking supplies. Future studies should capture more detailed information to understand the diversity and volume of safer smoking supplies distributed by SSPs.

## 5. Conclusions

The U.S. is currently experiencing its third decade of an overdose crisis. This study found SSPs implementing safer smoking pipe distribution had more participant engagement and naloxone distribution. To maximize their full individual and population-level health benefits, SSPs should be supported technically, legally, and financially to implement safer smoking supply distribution for their participants. Policymakers, federal agencies, and state and county health departments should be informed about the potential public health benefits of safer smoking supply distribution and address barriers to implementing this important service.

## Data availability

Presented data are available upon reasonable request from the corresponding author.

## Acknowledgements

The authors are deeply grateful to the syringe services programs and their representatives for participating in the National Survey of Syringe Services Program (NSSSP). The authors thank other members of the NSSSP Team—which includes partners at RTI International (Erica Browne) and the Centers for Disease Control and Prevention (Monica Adams, Aleta Christensen, Ryan Drab, Talia Pindyck) —who contributed substantially to the planning and implementation of the NSSSP. In addition to partnering with CDC and NASEN to administer the NSSSP, RTI collaborated with National Harm Reduction Coalition (NHRC) and National Alliance of State & Territorial AIDS Directors (NASTAD) to refine the instrument and promote the NSSSP to their networks of harm reductionists. The authors also thank other team members at RTI who supported survey administration and recruitment including Erin Erickson, Kathryn Greenwell, Terry Morris, and Abigail Rinderle.

