## Appendices for "Distributing Safer Smoking Pipes Increases Engagement with Harm Reduction Services in the United States: Findings from the National Survey of Syringe Services Programs"

**Appendix Methods**

**Multiple imputation**

There was substantial missingness in key variables (38.61%) with 23 unique missingness patterns (Appendix Table 1). The variable with the largest missingness was syringe services program (SSP) budget with roughly 34% missing, and the most common missing pattern was only missing SSP budget. Further, key characteristics were different by incomplete vs. complete case (Appendix Table 2). Therefore, we conducted multiple imputation to address missing data.

Multiple imputation assumes the data are missing completely at random (MCAR, the probability of missingness does not depend on unobserved or observed data) or missing at random (MAR, conditional on observed data, the probability of missingness does not depend on unobserved data). We assume here the data are MAR. We conducted multiple imputation using chained equations (MICE), a flexible model that allows for specification of missing variables based on its distribution (e.g. Gaussian, logistic, Poisson) (Van Buuren & Groothuis-Oudshoorn, 2011; White et al., 2011; Graham, 2009).

The imputation model included all analytic variables, including the outcomes and exposure. We used predictive mean matching for continuous variables and logistic and multinomial logistic for binary and categorical variables, respectively. To improve the imputation model, we included auxiliary variables that were associated with the missing variable(s) or the missingness of variable(s). Auxiliary variables are not of substantive interest in the analysis model, but their inclusion in the imputation model helps to improve the imputation of missing values by increasing power and by helping make the MAR assumption more plausible (Graham, 2009; White et al., 2011). We included the following auxiliary variables that were associated with the missing variables or the missingness of variables: number of weekly operating hours, number of funding sources, full-time staff employment, service areas (urban, suburban, rural), and SSP relationship with law enforcement (Very good, Somewhat good, Neither good nor bad, Somewhat poor, Very poor, Nonexistent).

We ran 50 imputed datasets including all analytic and auxiliary variables using the *mice* package in R. Based on summary statistics, histograms, and boxplots, the imputed data matched the observed data relatively well. Model convergence was adequate based on trace plots of imputed means and standard deviations for each imputed variable.

Based on prior recommendations to improve precision (Von Hippel, 2007; White et al., 2011), we restricted our analytic sample to observations with observed outcomes (n=429) – though we included these observations in the imputation model. We also ran two sensitivity models: (1) restricting to only complete cases (n=291) and (2) using the fully imputed dataset (n=471). We found higher precision (e.g. smaller confidence limit ratios [CLRs], Monte Carlo errors, and fraction of missing information) when we used MICE compared to complete case (Appendix Table 3). CLRs measure precision and represent the width of the 95% CI (i.e. upper limit divided by lower limit) (Poole, 2001). Analyses using the fully imputed dataset had lower precision (e.g. higher CLRs, Monte Carlo errors, and fraction of missing information) compared to when we restricted the sample to observations with complete information on outcomes (Appendix Table 4).

| **Appendix Table 1. Missing data patterns, NSSSP 2023 Survey** | | | | | | | | | |
| --- | --- | --- | --- | --- | --- | --- | --- | --- | --- |
| **Missing Pattern (R)** | **Pipe distribution** | **# of participant encounters** | **# of naloxone doses** | **SSP type** | **SSP urbanicity** | **SSP budget** | **Opioid overdose mortality** | **N** | **%** |
| 1 | O | O | O | O | O | O | O | 291 | 61.91 |
| 2 | O | O | O | O | O | M | O | 124 | 26.38 |
| 3 | M | M | M | O | M | M | M | 10 | 2.13 |
| 4 | O | O | M | O | O | M | O | 6 | 1.28 |
| 5 | O | O | O | O | M | O | M | 5 | 1.06 |
| 6 | O | M | M | O | O | M | O | 5 | 1.06 |
| 7 | O | O | M | O | O | O | O | 3 | 0.64 |
| 8 | O | M | O | O | O | M | O | 3 | 0.64 |
| 9 | O | O | O | O | M | M | M | 2 | 0.43 |
| 10 | O | O | O | M | M | M | M | 2 | 0.43 |
| 11 | O | M | O | O | O | O | O | 2 | 0.43 |
| 12 | O | M | M | O | O | O | O | 2 | 0.43 |
| 13 | O | M | M | M | O | M | O | 2 | 0.43 |
| 14 | M | O | O | O | M | O | M | 2 | 0.43 |
| 15 | M | O | M | O | M | M | M | 2 | 0.43 |
| 16 | M | M | M | M | M | M | M | 2 | 0.43 |
| 17 | O | O | O | O | O | O | M | 1 | 0.21 |
| 18 | O | O | O | M | O | O | O | 1 | 0.21 |
| 19 | O | O | O | M | O | M | O | 1 | 0.21 |
| 20 | O | M | O | O | M | O | M | 1 | 0.21 |
| 21 | O | M | M | O | M | O | M | 1 | 0.21 |
| 22 | O | M | M | O | M | M | M | 1 | 0.21 |
| 23 | M | M | M | O | O | M | O | 1 | 0.21 |
| Total missing  N (%) | 17 (3.61) | 30 (6.37) | 35 (7.43) | 8 (1.70) | 28 (5.94) | 161 (34.18) | 29 (6.16) | 470 | 100% |
| O = observed, M = missing. Other variables not included in this table were fully observed (census division). | | | | | | | | | |

| **Appendix Table 2. Descriptive statistics by incomplete vs. complete cases, NSSSP 2023 survey, N=471** | | | |
| --- | --- | --- | --- |
|  | Incomplete | Complete | Total |
|  | 179 (38.0%) | 291 (61.8%) | 471 (100.0%) |
| Smoking pipe distribution | 47.0 (28.8%) | 151.0 (51.9%) | 198.0 (43.6%) |
| Participant contacts | 821 [68 - 3,208] | 1,600 [500 - 4,901] | 1,400 [268 - 4,099] |
| Naloxone doses distributed | 1,000 [72 - 4,624] | 2,798 [998 - 7,740] | 2,154 [500 - 6,031] |
| SSP Type |  |  |  |
| CBO | 69.0 (40.1%) | 198.0 (68.0%) | 267.0 (57.7%) |
| DPH/HCO/Other | 103.0 (59.9%) | 93.0 (32.0%) | 196.0 (42.3%) |
| Urbanicity |  |  |  |
| Rural | 50.0 (32.9%) | 59.0 (20.3%) | 109.0 (24.6%) |
| Suburban | 71.0 (46.7%) | 140.0 (48.1%) | 211.0 (47.6%) |
| Urban | 31.0 (20.4%) | 92.0 (31.6%) | 123.0 (27.8%) |
| Total annual budget |  |  |  |
| Less than 149k | 7.0 (36.8%) | 149.0 (51.2%) | 156.0 (50.3%) |
| >150k | 12.0 (63.2%) | 142.0 (48.8%) | 154.0 (49.7%) |
| Opioid overdose death rate per 100,000 (2020) | 30.2 [24.2 - 43.3] | 30.8 [21.6 - 41.1] | 30.6 [22.7 - 41.1] |
| Census Divisions |  |  |  |
| New England | 13.0 (7.3%) | 21.0 (7.2%) | 34.0 (7.2%) |
| Middle Atlantic | 6.0 (3.4%) | 21.0 (7.2%) | 27.0 (5.7%) |
| East North Central | 14.0 (7.8%) | 55.0 (18.9%) | 69.0 (14.7%) |
| West North Central | 6.0 (3.4%) | 16.0 (5.5%) | 22.0 (4.7%) |
| South Atlantic | 26.0 (14.5%) | 56.0 (19.2%) | 82.0 (17.4%) |
| East South Central | 21.0 (11.7%) | 17.0 (5.8%) | 38.0 (8.1%) |
| West South Central | 2.0 (1.1%) | 14.0 (4.8%) | 16.0 (3.4%) |
| Mountain | 68.0 (38.0%) | 25.0 (8.6%) | 93.0 (19.8%) |
| Pacific | 23.0 (12.8%) | 66.0 (22.7%) | 89.0 (18.9%) |

| **Appendix Table 3. Sensitivity analyses comparing model diagnostics between complete case and MICE** | | | | |
| --- | --- | --- | --- | --- |
|  | **Participant encounters** | | **Naloxone doses distributed** | |
|  | Complete case (n=291) | MICE excluding imputed outcomes (n=429) | Complete case (n=291) | MICE excluding imputed outcomes (n=429) |
| Rate ratio | 1.41 | 1.49 | 1.46 | 1.21 |
| 95% Confidence interval | 1.00-1.99 | 1.09-2.02 | 1.07-1.97 | 0.89-1.65 |
| Confidence limit ratio | 1.99 | 1.85 | 1.84 | 1.85 |

| **Appendix Table 4. Comparing model diagnostics between MICE models** | | | | |
| --- | --- | --- | --- | --- |
|  | **Participant encounters** | | **Naloxone doses distributed** | |
|  | Excluding imputed outcomes (n=429) | Including imputed outcomes (n=471) | Excluding imputed outcomes (n=429) | Including imputed outcomes (n=471) |
| Rate ratio | 1.485 | 1.509 | 1.213 | 1.241 |
| Standard error | 0.156 | 0.158 | 0.155 | 0.156 |
| 95% Confidence Interval | 1.09-2.02 | 1.11-2.06 | 0.89-1.65 | 0.91-1.69 |
| Confidence limit ratio | 1.85 | 1.86 | 1.85 | 1.86 |
| Monte Carlo error | 0.00539 | 0.00839 | 0.00629 | 0.00873 |
| Fraction of missing information | 0.0657 | 0.15 | 0.0887 | 0.164 |
